# Intermittent Lactobacilli-containing Vaginal Probiotic or Metronidazole Use to Prevent Bacterial Vaginosis Recurrence: Safety and Preliminary Efficacy by Microscopy and Sequencing

**DOI:** 10.1101/19001156

**Authors:** Janneke H.H.M. van de Wijgert, Marijn C. Verwijs, Stephen K. Agaba, Christina Bronowski, Lambert Mwambarangwe, Mireille Uwineza, Elke Lievens, Adrien Nivoliez, Jacques Ravel, Alistair C. Darby

**Author notes:** Correspondence and requests for materials should be addressed to JvdW;). Janneke H.H.M. van de Wijgert and Marijn C. Verwijs contributed equally to this work.

## Abstract

Bacterial vaginosis (BV) is associated with HIV acquisition and adverse pregnancy outcomes. Recurrence after metronidazole treatment is high. HIV-negative, non-pregnant Rwandan BV patients were randomized to four groups (n=17/group) after seven-day oral metronidazole treatment: behavioral counseling only (control), or counseling plus intermittent use of oral metronidazole, Ecologic Femi+ vaginal capsule (containing multiple *Lactobacillus* and one *Bifidobacterium* species), or Gynophilus LP vaginal tablet (*L. rhamnosus* 35) for two months. Vaginal microbiota assessments at all visits included Gram stain Nugent scoring and 16S rRNA gene qPCR and HiSeq sequencing. All interventions were safe. BV (Nugent 7-10) incidence was 10.18 per person-year at risk in the control group, and lower in the metronidazole (1.41/person-year; p=0.004), Ecologic Femi+ (3.58/person-year; p=0.043), and Gynophilus LP groups (5.36/person-year; p=0.220). In mixed effects models adjusted for hormonal contraception/pregnancy, sexual risk-taking, and age, metronidazole and Ecologic Femi+ users, each compared to controls, had higher *Lactobacillus* and lower BV-anaerobes concentrations and/or relative abundances, and were less likely to have a dysbiotic vaginal microbiota type by sequencing. Inter-individual variability was high and effects disappeared soon after intervention cessation. Lactobacilli-based vaginal probiotics warrant further evaluation because, in contrast to antibiotics, they are not expected to negatively affect microbiota or cause antimicrobial resistance.

## INTRODUCTION

Most women have a vaginal microbiota (VMB) that consists predominantly of lactobacilli^1^. The most common type of vaginal dysbiosis is bacterial vaginosis (BV), characterized by a decrease in lactobacilli and increase in other anaerobes, which affects 30-40% of women worldwide^2^. Other types of bacterial dysbiosis, vulvovaginal candidiasis, and *Trichomonas vaginalis* (TV) are also common. These conditions are associated with vaginal inflammation, thereby increasing the risk of HIV acquisition^3^. They are also associated with pelvic inflammatory disease, infertility, and adverse pregnancy outcomes^2^.

The majority of women seeking care for vaginal symptoms receive antibiotic or antifungal treatment empirically or syndromically without any diagnostic testing^4^. In some specialized clinics, women might be offered limited diagnostic testing, such as vaginal pH determination and/or wet mount microscopy. In research settings, BV is typically diagnosed by the Amsel criteria or Nugent scoring^5,6^, with the latter currently being considered the gold standard: Gram-stained vaginal smears are scored based on microscopic visualization of three bacterial morphotypes with a score of 0-3 considered normal, 4-6 intermediate, and 7-10 BV regardless of symptoms. In the last 15 years, molecular methods have become more widely available, and have been applied to the VMB, although mostly in descriptive studies to date^1^.

Evidence is mounting that ‘microbiological-BV’ (by Nugent scoring or molecular methods) can cause long-term adverse outcomes in the presence or absence of vaginal symptoms^2^. BV is notoriously difficult to treat^7–9^. About 60-80% of patients are cured after a course of oral or vaginal metronidazole or clindamycin, but recurrence rates are high^7^. Therapies to restore and maintain an optimal lactobacilli-dominated VMB after antibiotic treatment are not standard practice, but some clinicians in Europe and North America recommend twice weekly 0.75% metronidazole vaginal gel for 4-6 months to lower the risk of BV recurrence^7^. This recommendation was tested in a randomized controlled trial in the USA, which showed a statistically significant reduction in BV recurrence (34% by Nugent scoring)^10^. In addition, two African trials evaluating oral (2g monthly) and vaginal metronidazole (five nights every three months) for BV prevention showed significant beneficial effects, but the effects were modest, most likely due to infrequent dosing^11,12^.

Probiotic lactobacilli may be able to restore and maintain a lactobacilli-dominated VMB, may be better able to prevent or disrupt BV-associated biofilms than antibiotics, and can likely be used safely for long periods without the risk of causing antimicrobial resistance^13^. Lactobacilli-containing vaginal probiotic clinical trials to date have shown mixed results, but eight of the 12 trials showed sufficiently promising efficacy for BV prevention to warrant further investigation^13–24^. Some trials used commercially available probiotic strains (mostly derived from the gut or fermented foods) and others vaginal strains isolated from healthy women^25^, but efficacy signals were similar for the two probiotic strain categories^13–24^. None of the trials reported major safety concerns or colonization beyond the dosing period. Because ‘natural’ strains do not seem to outperform commercially available strains, and for pragmatic reasons, we chose to evaluate two vaginal probiotics that are currently on the market. Our aim was to assess their safety and impact on the VMB in much more detail than previous trials had done, and to develop data analytic methods to enable use of high-dimensional 16S rRNA gene sequencing data for this purpose. The two probiotics that we evaluated were Ecologic Femi+ vaginal capsule (EF+; Winclove Probiotics, Amsterdam, The Netherlands) and Gynophilus LP vaginal tablet (GynLP; Biose, Arpajon-sur-Cère, France). EF+ contains multiple lyophilized bacteria (*Bifidobacterium bifidum* W28, *Lactobacillus acidophilus* W70, *L. helveticus* W74, *L. brevis* W63, *L. plantarum* W21, and *L. salivarius* W24) in a total dose of 1.5×10^9^ colony forming units (CFU). GynLP contains 1.6×10^9^ CFU of Lcr Regenerans, a culture of the *L. rhamnosus* 35 (Lcr35) strain. The tablet disintegrates in the vagina after forming a gel to release Lcr35. Gynophilus (the same active ingredient as GynLP but a different formulation) had shown promise in preventing BV recurrence in a previous trial, but EF+ had not previously been studied for this indication^15^.

## METHODS

From June 2015 until February 2016, we conducted a randomized clinical trial in Kigali, Rwanda, to evaluate intermittent use of the above-mentioned vaginal probiotics as well as oral metronidazole (Tricozole, Laboratory & Allied ltd, Nairobi, Kenya) to prevent the recurrence of microbiological-BV after metronidazole treatment. The trial was a pilot trial with a modest sample size (N=68) at the request of the funder.

### Eligibility

Women were eligible for screening if they were aged 18-45, in good overall physical and mental health, and at high urogenital infection risk defined as having had more than one sex partner in the last 12 months or having been treated for a sexually transmitted infection and/or BV in the last 12 months. They were eligible for enrollment if they were confirmed HIV-negative and non-pregnant, were diagnosed with BV (by Nugent score and/or Amsel criteria) and/or TV (by wet mount and/or culture), and were cured after seven days of oral metronidazole treatment (500 mg twice per day)^5^. Cure was defined as no BV by Amsel criteria and no TV by wet mount^6^. We did not use Nugent scores and TV culture as tests-of-cure to allow for same day enrollment but results became available after enrollment. At enrollment, 51/68 women were BV-negative by Amsel and Nugent criteria, 17/68 women were BV-negative by Amsel but BV-positive by Nugent criteria, and all women were TV-negative by both wet mount and culture and free of symptomatic vulvovaginal candidiasis, urinary tract infection, syphilis, and clinician-observed genital abnormalities or vaginal discharge. We did not exclude women with gonorrhea and/or chlamydia because the local testing turn-around time was slow. Positive herpes simplex type 2 serology was not a reason for exclusion. Additional exclusion criteria applied but these were rare (Figure 1, Supplement 1).

**Figure 1.**
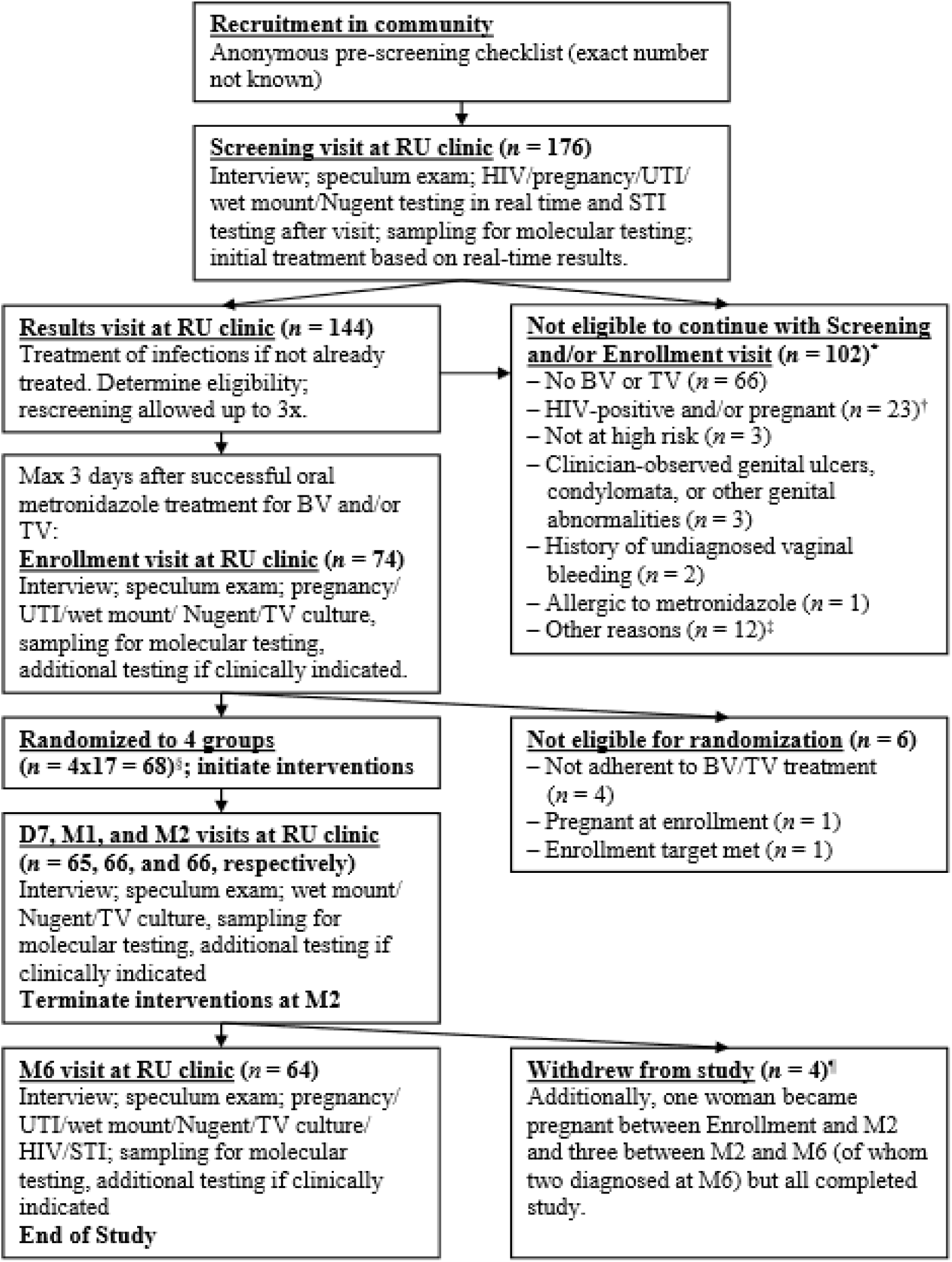
Flow diagram of numbers of women seen and study procedures at each visit. Abbreviations: BV, bacterial vaginosis; D7, day 7 visit; M1/2/6, month 1/2/6 visit; RU, Rinda Ubuzima; STI, sexually transmitted infection; TV, *Trichomonas vaginalis*; UTI, urinary tract infection. *Totals to 110 reasons among 102 women because there could be more than one reason per woman. †No speculum exam performed; molecular testing of self-sampled vaginal swabs. ‡Reasons: outside of metronidazole treatment window (n=5), enrollment target already met (n=4), has a mental disorder (n=1), did not complete screening procedures and was subsequently lost to follow=up (n=1), withdrew consent during the Screening visit because she thought the reimbursement was too low (n=1). §Three women in each randomization group were selected for self-sampling every other day during the first month of follow-up. ¶Reasons: moved away from Kigali (n=2), lost interest because symptoms resolved (n=1), and was verbally harassed by partner and sister about study participation (n=1).

### Randomization groups and visit schedule

Women were randomized to four groups (17 women per group) within three days of completing oral metronidazole treatment: 1) behavioral counseling only (control group); 2) counseling plus 500 mg oral metronidazole twice weekly for two months; 3) counseling plus EF+ vaginal capsule once per day for the first five days followed by thrice weekly for a total of two months; and 4) counseling plus GynLP once every four days for two months. Participants applied the first dose of their intervention under direct observation at the enrollment visit, and returned to the clinic after seven days (D7), one month (M1), two months (M2; cessation of product use), and six months (M6). They were allowed to cease vaginal product use temporarily during menstruation. Product adherence was assessed by review of diary cards, used and unused products, and by a self-rating adherence scale. Symptomatic BV, vulvovaginal candidiasis, and urinary tract infections, and laboratory-confirmed sexually transmitted infections, diagnosed during follow-up were treated using standard oral therapies, and women were urged to continue their interventions during treatment. Visit procedures are summarized in Figure 1 and described in Supplement 1.

### Diagnostic testing

Women were tested for BV, TV, and vulvovaginal candidiasis at each visit. BV was diagnosed by Gram stain Nugent scoring^5^ and Amsel criteria^6^; the vaginal pH was measured by pressing a pH paper strip (pH range 3.6 – 6.1 with 0.3 increments; Machery-Nagel, Düren, Germany) against the vaginal wall. TV was diagnosed when motile trichomonads were observed on wet mount and by InPouch culture (Biomed Diagnostics, White City, OR, USA). Vulvovaginal candidiasis was diagnosed when budding yeasts and/or (pseudo)hyphae were observed on wet mount. All other diagnostic tests (Supplement 1) were only done at screening, M6, and when judged clinically necessary by the physician, with the exception of pregnancy and urinalysis tests, which were repeated at enrollment prior to randomization.

### 16S rRNA gene sequencing and qPCR

We collected vaginal swabs at 639 time points, including study visits of all 68 women and self-sampled swabs from 12 women (three per group) who had been asked to self-sample at home every Monday, Wednesday, and Friday during the first month. Self-sampled swabs were processed but were not included in analyses unless stated. DNA was extracted from one swab per time point per woman. Briefly, DNA was extracted using a combination of lysozyme lysis, Qiagen DNeasy Blood and Tissue kit (Qiagen, Manchester, UK), and bead-beating procedures. Purified DNA underwent two PCR rounds: amplification of the 16S rRNA gene V3-V4 region and dual-index barcoding allowing multiplexing of up to 384 samples. Samples were sequenced on an Illumina HiSeq 2500 instrument (Illumina, San Diego, CA, USA), run in rapid mode, 2×300bp using a 250PE and 50PE kit. All samples collected from the same participant were included in the same run. Negative and positive controls (ZymoBiomics Microbial Community DNA standard; Zymo Research Corp, Irvine, USA) were incorporated throughout. The panbacterial 16S rRNA gene copy concentrations of physician-collected samples of enrolled women with valid sequencing results (N=393) were determined by BactQuant qPCR assay as previously described^26^.

### Molecular data processing

Reads were demultiplexed, and primer sequences removed using Cutadapt v1.2.1^27^. Error correction, dereplication, denoising, merging, removal of chimeras, and taxonomic assignment were performed in DADA2 v1.4.0 using Silva v128 as the reference database (Supplement 1)^28,29^. Further data processing included removal of rare, non-bacterial, and contaminant amplicon sequence variants (ASVs), and identification of vaginal and probiotic sequences that are not included in the Silva database using the NCBI *Needleman-Wunsch Global Align Nucleotide Sequences* function^15^ (Supplement 1). Silva-based taxonomic assignments of non-minority ASVs were double-checked using the NCBI *Microbial Nucleotide BLAST* function, using the non-redundant V3-V4 version of the Vaginal 16S rDNA Reference Database as a tiebreaker^30,31^. We rarefied at 1,111 reads using the *GUniFrac 1*.*0* package in R^32^. This yielded 401 unique ASVs in 629 samples, mapping to species (n=255; 63.6%), genus (n=116; 28.9%), or higher taxonomic level (n=30; 7.5%). Concentrations in cells/μl per ASV per sample were estimated by multiplying the ASV-specific copy-normalized relative abundance by the sample-specific 16S rRNA gene copies concentration^33,34^. Heatmaps of the 20 most common ASVs by sample are shown in Figure S1A for relative abundances and Figure S1B for concentrations. VMB data reduction was required for molecular efficacy analyses, and was done in three different ways. First, the Simpson diversity index (1-D) was calculated for each sample. Second, each ASV was assigned to one of four ‘bacterial groups’ based on the published literature (Supplement 2): 1) lactobacilli (all species combined, but with subcategorization into EF+ strains, the GynLP strain, and ‘natural lactobacilli’ in some analyses); 2) BV-anaerobes (anaerobic bacteria that have consistently been associated with BV); 3) pathobionts (bacteria that are considered to have higher intrinsic pathogenicity than BV-anaerobes and have not consistently been associated with BV^35^); and 4) ‘other bacteria’ (a rest group, consisting mostly of skin bacteria and *Bifidobacterium* species). Within each sample, read counts of ASVs belonging to the same bacterial group were summed. This resulted in four relative abundances (one for each bacterial group) per sample, which sum to 1.0 for each sample. Third, we used hierarchical clustering based on Euclidean distance to pool samples into eight mutually exclusive VMB types (each sample was assigned to only one VMB type): 1) *L. iners*-dominated (Li, n=247 samples); 2) *L. crispatus*-dominated (Lcr, n=17); 3) other lactobacilli-dominated (Lo, n=28); 4) lactobacilli and anaerobes (LA, n=86); 5) polybacterial *Gardnerella vaginalis*-containing (BV_GV, n=138), 6) polybacterial but low-*G. vaginalis* (BV_noGV, n=23), 7) *G. vaginalis*-dominated (GV, n=41); and 8) pathobionts-containing (PB, n=49) (Supplement 1).

### Downstream statistical analysis

Adverse events were coded using the Medical Dictionary for Regulatory Activities v19.1 (McLean, VA, USA). Data were analyzed using STATA v13.0 (StataCorp, College Station, TX, USA). Most statistical comparisons were between randomization groups (each intervention group compared to the control group) at screening, enrollment, and longitudinally over time. For cross-sectional analyses, we used Fisher’s exact test for binary and categorical variables, and Kruskal-Wallis and Mann Whitney U tests for continuous variables. For longitudinal analyses, we used incidence rates, incidence rate ratios, and mixed effects models. We conducted intent-to-treat (ITT) analyses, and modified ITT analyses limited to women who were BV-negative by both Amsel and Nugent criteria at the time of randomization (n=51 of 68). All mixed models included one VMB endpoint at a time as the outcome, participant identification number as the random effect, and randomization group (an indicator variable with the control group as the reference group) as the main fixed effect. Models included samples collected during the intervention period only (including self-sampled time points), and adjusted for covariates that were associated with VMB composition in mixed effects models at p<0.05.

### Ethical statement

All participants provided written informed consent. The study was conducted in accordance with the Declaration of Helsinki, sponsored by the University of Liverpool, approved by the Rwanda National Ethics Committee and the University of Liverpool Research Ethics Subcommittee for Physical Interventions, and registered on ClinicalTrials.gov (NCT02459665).

### Data availability

All data will be made available via the University of Liverpool Data Research Catalogue (https://datacat.liverpool.ac.uk/) with publication.

## RESULTS

### Participant disposition

Of the 68 randomized women, only four did not complete the trial (Figure 1), resulting in 29.93 person-years of data. The median age was 31 (interquartile range (IQR) 27-35) (Table 1, Table S1). The majority of women (92.6%) had exchanged sex for money or goods, and had had a median of five (IQR 2-18) sex partners, in the past month. All but three women used condoms, but mostly inconsistently. Two-thirds of the women (61.8%) were using hormonal contraception, and four women became pregnant during the trial. Short course metronidazole/tinidazole use for other indications during the intervention period was evenly distributed among randomization groups (Table S2: n=1-3 per group; p=0.688), as was short course use of other oral antibiotics (Table S2: n=2-4 per group; p=0.781). Furthermore, these other antibiotics did not impact lactobacilli and BV-anaerobes concentrations significantly (Table S2, Figure S2). No antifungals were used. Most women were adherent with their study product >90% of the time, but this percentage was non-significantly lower in the GynLP group (68.8%) than in the metronidazole (82.4%; Fisher’s exact p=0.438) and EF+ groups (88.2%, Fisher’s exact p=0.225; Table S3).

**Table 1.** Baseline characteristics.

| **Sociodemographic characteristics at Scr/Enr** | **Screened (n = 175)** | **Enrolled (n = 68)** | **Controls (n = 17)** | **Metro (n = 17)** | **EF+ (n = 17)** | **GynLP (n = 17)** | **p\*** |
| --- | --- | --- | --- | --- | --- | --- | --- |
| Age in years, median (IQR) | 30 (27 – 34) | 31 (27 – 35) | 29 (24 – 36) | 30 (27 – 34) | 33 (28 – 35) | 30 (27 – 35) | 0.563 |
| Sex partners last mo, median (IQR) | 5 (2 – 16) | 5 (2 – 18) | 5 (3 – 20) | 5 (2 – 10) | 3 (2 – 15) | 3 (2 – 20) | 0.624 |
| Condom during last sex act, n (%)<sup>†</sup> | 95 (54.6) | 36 (52.9) | 11 (64.7) | 8 (47.1) | 6 (35.3) | 11 (64.7) | 0.279 |
| Currently using contraception, n (%): |  |  |  |  |  |  |  |
| - None | 69 (40.2) | 24 (35.3) | 6 (35.3) | 6 (35.3) | 4 (23.5) | 8 (47.1) | 0.750 |
| - Hormonal contraception | 99 (57.6) | 42<sup>‡</sup> (61.8) | 10 (58.8) | 11 (64.7) | 12 (70.6) | 9 (52.9) |  |
| - Copper IUD | 4 (2.3) | 2 (2.9) | 1 (5.9) | 0 | 1 (5.9) | 0 |  |
| **Laboratory results at Scr** | **Screened (n = 173)** | **Enrolled (n = 68)** | **Controls (n = 17)** | **Metro (n = 17)** | **EF+ (n = 17)** | **GynLP (n = 17)** | **p\*** |
| Nugent score, mean (95% CI)<sup>†</sup> | 4.7 (4.1 – 5.3) | 7.4 (6.8 – 8.0) | 7.6 (6.5 – 8.6) | 6.8 (5.3 – 8.3) | 7.1 (5.5 – 8.6) | 8.2 (7.4 – 9.0) | 0.333 |
| Nugent categories, n (%)<sup>†</sup> |  |  |  |  |  |  |  |
| - 0-3 | 55 (38.2)<sup>§</sup> | 5 (7.5) | 1 (5.9) | 2 (11.8) | 2 (12.5) | 0 | 0.075 |
| - 4-6 | 20 (13.9) | 6 (9.0) | 0 | 4 (23.5) | 0 | 2 (11.8) |  |
| - 7-10 | 69 (47.9) | 56 (83.5) | 16 (94.1) | 11 (64.7) | 14 (87.5) | 15 (88.2) |  |
| BV by Amsel criteria, n (%) | 30 (20.6) | 20 (29.4) | 4 (23.5) | 6 (35.5) | 3 (17.7) | 7 (41.2) | 0.486 |
| Candida on wet mount, n (%) | 14 (9.6) | 6 (8.8) | 1 (5.9) | 2 (11.8) | 3 (17.7) | 0 | 0.493 |
| TV on wet mount, n (%) | 9 (6.2) | 6 (8.8) | 3 (17.7) | 1 (5.9) | 1 (5.9) | 1 (5.9) | 0.707 |
| TV by InPouch culture, n (%) | 17 (11.8)<sup>§</sup> | 11 (16.4) | 3 (18.8) | 1 (5.9) | 5 (29.4) | 2 (11.8) | 0.282 |
| UTI by dipstick, n (%) | 33 (19.1) | 17 (25.0) | 4 (23.5) | 6 (35.3) | 4 (23.5) | 3 (17.7) | 0.760 |
| CT by PCR, n (%) | 30 (20.8)<sup>§</sup> | 20 (29.4) | 5 (29.4) | 7 (41.2) | 3 (17.7) | 5 (29.4) | 0.560 |
| NG by PCR, n (%) | 18 (12.5)<sup>§</sup> | 13 (19.1) | 5 (29.4) | 4 (23.5) | 2 (11.8) | 2 (11.8) | 0.555 |
| HIV serology positive, n (%) | 17 (9.8) | 0 | 0 | 0 | 0 | 0 | NA |
| HSV2 serology positive, n (%) | 117 (67.6) | 44 (64.7) | 9 (52.9) | 12 (70.6) | 11 (64.7) | 12 (70.6) | 0.780 |
| Syphilis serology positive, n (%) | 13 (7.5) | 4 (5.9) | 0 | 1 (5.9) | 0 | 3 (17.7) | 0.182 |
| Pregnancy test positive, n (%) | 7 (4.1) | 0 | 0 | 0 | 0 | 0 | NA |
| **Laboratory results at Enr\*\*** | **Screened (n = 176)** | **Enrolled (n = 68)** | **Controls (n = 17)** | **Metro (n = 17)** | **EF+ (n = 17)** | **GynLP (n = 17)** | **p\*** |
| Nugent score, mean (95% CI)<sup>†</sup> | NA | 3.1 (2.2 – 3.9) | 3 (1.2 – 4.8) | 3.3 (1.4 – 5.2) | 1.7 (0.1 – 3.3) | 4.3 (2.6 – 6.0) | 0.187 |
| Nugent categories, n (%)<sup>†</sup> |  |  |  |  |  |  |  |
| - 0-3 | NA | 36 (54.6) | 8 (53.3) | 9 (52.9) | 13 (76.5) | 6 (35.3) | 0.149 |
| - 4-6 | NA | 13 (19.7) | 5 (33.3) | 2 (11.8) | 1 (5.9) | 5 (29.4) |  |
| - 7-10 | NA | 17 (25.8) | 2 (13.3) | 6 (35.3) | 3 (17.7) | 6 (35.3) |  |
| BV by Amsel criteria, n (%) | NA | 0 | 0 | 0 | 0 | 0 | NA |
| Candida on wet mount, n (%) | NA | 4 (5.9) | 2 (11.8) | 1 (5.9) | 0 | 1 (5.9) | 0.897 |
| TV by wet mount/culture, n (%) | NA | 0 | 0 | 0 | 0 | 0 | NA |
| **VMB outcomes at Enr** | **Screened (n = 176)** | **Enrolled (n = 67)** | **Controls (n = 17)** | **Metro (n = 17)** | **EF+ (n = 17)** | **GynLP (n = 16)** | **p\*** |
| Richness, mean (95% CI) | NA | 12.8 (10.7 – 14.8) | 17.8 (12.1 – 23.5) | 12.1 (8.7 – 15.5) | 7.5<sup>‡‡</sup> (4.9 – 10.2) | 13.7 (10.3 – 17.1) | 0.003 |
| Simpson diversity index, mean (95% CI) | NA | 0.31 (0.25 – 0.38) | 0.38 (0.21 – 0.54) | 0.35 (0.20 – 0.50) | 0.15<sup>§§</sup> (0.05 – 0.25) | 0.38 (0.25 – 0.50) | 0.022 |
| Total bacteria concentration in log<sub>10</sub>/μL, mean (95% CI)<sup>††</sup> | NA | 5.85 (5.66 – 6.04) | 5.75 (5.30 – 6.20) | 5.79 (5.39 – 6.19) | 5.54 (5.28 – 5.80) | 6.34<sup>‡‡</sup> (5.95 – 6.73) | 0.019 |
| Total *Lactobacillus* concentration in log<sub>10</sub>/μL, mean (95% CI)<sup>††</sup> | NA | 5.56 (5.34 – 5.78) | 5.22 (4.63 – 5.82) | 5.59 (5.20 – 5.97) | 5.46 (5.19 – 5.73) | 5.97 (5.44 – 6.49) | 0.092 |
| Total BV-anaerobes concentration in log<sub>10</sub>/μL, mean (95% CI)<sup>††</sup> | NA | 4.55 (4.15 – 4.95) | 4.78 (4.17 – 5.39) | 4.50 (3.77 – 5.24) | 3.36<sup>‡‡</sup> (2.43 – 4.29) | 5.62<sup>‡‡</sup> (4.99 – 6.25) | 0.002 |
| Total pathobionts concentration in log<sub>10</sub>/μL, mean (95% CI)<sup>††</sup> | NA | 2.01 (1.48 – 2.54) | 2.36 (1.28 – 3.44) | 2.08 (1.00 – 3.17) | 1.34 (0.29 – 2.39) | 2.30 (0.98 – 3.62) | 0.447 |
| Total other bacteria concentration in log<sub>10</sub>/μL, mean (95% CI)<sup>††</sup> | NA | 1.46 (1.01 – 1.92) | 1.80 (0.84 – 2.76) | 1.30 (0.36 – 2.24) | 0.57 (-0.10 – 1.24) | 2.20 (1.07 – 3.34) | 0.066 |
| VMB types, n (%): |  |  |  |  |  |  |  |
| - Li | NA | 35 (52.2) | 7 (41.2) | 9 (52.9) | 14 (82.4) | 5 (31.3) | 0.048 |
| - Lcr | NA | 0 | 0 | 0 | 0 | 0 |  |
| - Lo | NA | 2 (3.0) | 1 (5.9) | 1 (5.9) | 0 | 0 |  |
| - LA | NA | 18 (26.9) | 3 (17.7) | 5 (29.4) | 2 (11.8) | 8 (50.0) |  |
| - BV_GV |  | 2 (3.0) | 1 (5.9) | 0 | 0 | 1 (5.9) |  |
| - BV_noGV |  | 0 | 0 | 0 | 0 | 0 |  |
| - GV |  | 4 (6.0) | 2 (11.8) | 0 | 0 | 2 (12.5) |  |
| - PB |  | 6 (9.0) | 3 (17.7) | 2 (11.8) | 1 (5.9) | 0 |  |

### VMB compositions at baseline

By design, all randomized women at screening had BV (by Nugent and/or Amsel criteria) and/or TV (by wet mount and/or culture): 82.4% had BV alone, 14.7% had BV and TV, and 2.9% had TV alone. Therefore, as expected, most women had dysbiotic VMB types by 16S rRNA gene sequencing at screening: BV_GV (41.8%), LA (17.9%), BV_noGV (11.9%), GV (11.9%), and PB (1.5%). However, 14.9% had a lactobacilli-dominated Li VMB type, of whom 60% were TV-negative, and these women would not have needed metronidazole treatment. Also by design, all randomized women were BV-negative (by Amsel criteria) and TV-negative (by wet mount and culture) at the time of randomization. However, almost half of the women (44.8%) were not lactobacilli-dominated by sequencing at the time of randomization, and none of them were *L. crispatus*-dominated (which is considered the most optimal VMB state): their VMB types were Li (52.2%), Lo (3.0%), LA (26.9%), PB (9.0%), GV (6.0%), and BV_GV (3.0%) (Table 1).

At randomization, the mean total bacterial concentration ranged from 5.54-6.34 log_10_ cells/μl in the four randomization groups, and these ranges were 5.22-5.97 for lactobacilli, 3.36-5.62 for BV-anaerobes, 1.34-2.36 for pathobionts, and 0.57-2.20 for ‘other bacteria’ (Table S4; relative abundance data in Table S5). The mean richness ranged from 7.5-17.8, and the mean Simpson diversity from 0.15-0.38. Randomization did not completely balance the baseline VMB compositions of the four groups: the concentrations total bacteria and BV-anaerobes were higher in the GynLP group than the control group (Mann Whitney U test p<0.05), and the BV-anaerobes concentration (p<0.01) and Simpson diversity (p<0.05) were lower in the EF+ group than the control group (Table 1).

Variables that were associated with at least one VMB composition variable in unadjusted mixed effects models (Table S6) included currently using hormonal contraception or being pregnant (associated with a higher pathobionts concentration), above-average sexual risk taking based on reported numbers of partners and condom use (higher pathobionts concentration), aged 30 years or older (lower BV-anaerobes and pathobionts concentrations), and managing menses with a sanitary pad compared to other methods (higher Nugent score). Reporting current urogenital symptoms was also associated with VMB composition but this likely represents reverse causality. We could not exclude women with ongoing chlamydia and/or gonorrhea infections at randomization because the turn-around time of diagnostic testing was slow, but the VMB compositions after metronidazole treatment of women with and without ongoing chlamydia and/or gonorrhea infection were similar (Figure S2). As mentioned earlier, short course antibiotic use that was not part of the study interventions was not associated with VMB composition either (Tables S2 and S6; Figure S2).

### Safety

Two serious adverse events occurred but these were judged not to be related to study participation: one woman in the oral metronidazole group was hospitalized for typhoid fever and one woman in the EF+ group for malaria during pregnancy. Both events occurred after the intervention period and both women recovered completely. Two women reported non-serious social harms that were judged related to study participation. One woman in the control group was beaten by her partner because she engaged in self-sampling; she withdrew from self-sampling but continued participation in the study. One woman in the GynLP group suffered verbal harassment from her partner and her sister for taking part in the study and elected to withdraw.

During the intervention period, urogenital symptoms were reported by 13.4% of participants (genital itching, burning, and pain during sex but no vaginal discharge) with no differences between groups, and only two speculum exam and no bimanual exam findings were reported by the physician (Table 2). After product cessation, urogenital symptom reporting was similar compared to the intervention period (10.8%), but the number of speculum exam findings increased (32.8%), likely reflecting the high urogenital infection incidence in this cohort. A total of 41 adverse events were spontaneously reported between enrollment and M6. All of them were judged definitely not or unlikely to be related to trial participation, and they were evenly distributed among groups.

**Table 2.** Safety endpoints.

| **Patient-reported symptoms and clinician-observed signs at Scr and Enr** | **Total**<br>**(n = 68)** | **Controls**<br>**(n = 17)** | **Metro**<br>**(n = 17)** | **EF+**<br>**(n = 17)** | **GynLP**<br>**(n = 17)** | **p\*** |
| --- | --- | --- | --- | --- | --- | --- |
| Any (current or in last 2 weeks) urogenital symptoms at Scr, n (%) | 49 (72.1)<sup>†</sup> | 11 (64.7) | 13 (76.5) | 13 (76.5) | 12 (70.6) | 0.933 |
| Any (current) urogenital symptoms at Enr, n (%) | 0 | 0 | 0 | 0 | 0 | NA |
| Any abnormal speculum exam findings at Scr, n (%)<sup>‡</sup> | 3 (4.4)<sup>§</sup> | 1 (5.9) | 1 (5.9) | 0 | 1 (5.9) | 1.00 |
| Any abnormal speculum exam findings at Enr, n (%)<sup>‡</sup> | 1 (1.5)<sup>¶</sup> | 0 | 0 | 0 | 1 (5.9) | 1.00 |
| **AEs – Structurally assessed between Enr and M2 (during product use)** |  |  |  |  |  |  |
| Any current urogenital symptoms, n (%) | 9 (13.4)<sup>**</sup> | 2 (11.8) | 2 (11.8) | 2 (11.8) | 3 (18.8) | 0.894 |
| Any abnormal speculum exam findings, n (%) | 2 (3.0)<sup>††</sup> | 0 | 0 | 0 | 2 (12.5) | 0.054 |
| Any abnormal bimanual exam findings n (%) | 0 | 0 | 0 | 0 | 0 | NA |
| **AEs – Structurally assessed between M2 and M6 (after product cessation)** |  |  |  |  |  |  |
| Any current urogenital symptoms, n (%) | 7 (10.8)<sup>‡‡</sup> | 3 (17.7) | 1 (5.9) | 1 (6.3) | 2 (13.3) | 0.700 |
| Any abnormal speculum exam findings, n (%) | 21 (32.8)<sup>§§</sup> | 6 (35.3) | 5 (31.3) | 6 (37.5) | 4 (26.7) | 0.951 |
| Any abnormal bimanual exam findings n (%) | 1 (1.6)<sup>¶¶</sup> | 0 | 0 | 0 | 1 (6.7) | 0.234 |
| **AEs – Not structurally assessed** | **Total (N)** | **Controls**<br>**(N)** | **Metro (N)** | **EF+ (N)** | **GynLP**<br>**(N)** | **p\*** |
| Number of women with reported AEs | 27 | 8 | 4 | 8 | 7 | 0.439 |
| Total number reported AEs between Enr-M6 | 41 | 13 | 6 | 9 | 13 | 0.324 |
| Total number reported AEs between Enr-M2 | 32 | 12 | 4 | 5 | 11 | NA |
| Total number reported AEs between M2-M6<sup>***</sup> | 9 | 1 | 2 | 4 | 2 | NA |
| **AEs – Not structurally assessed** | **Total AEs**<br>**(n = 41)** | **Controls**<br>**(n = 13)** | **Metro**<br>**(n = 6)** | **EF+**<br>**(n = 9)** | **GynLP**<br>**(n = 13)** | **p\*** |
| Severity of reported AEs, n (%): |  |  |  |  |  |  |
| - Mild | 2 (4.9) | 0 | 0 | 1 (11.1) | 1 (7.7) | NA |
| - Moderate | 36 (87.8) | 13 (100) | 5 (83.3) | 6 (66.7) | 12 (92.3) |  |
| - Severe | 3 (7.3) | 0 | 1 (16.7) | 2 (22.2) | 0 |  |
| - Life-threatening | 0 | 0 | 0 | 0 | 0 |  |
| Deemed related to study by physician, n (%): |  |  |  |  |  |  |
| - Definitely not related | 10 (24.4) | 3 (23.1) | 2 (33.3) | 3 (33.3) | 2 (15.4) | NA |
| - Unlikely | 31 (75.6) | 10 (76.9) | 4 (66.7) | 6 (66.7) | 11 (84.6) |  |
| - Possible/probable/definitely related | 0 | 0 | 0 | 0 | 0 |  |
| Outcome of reported AE, n (%): |  |  |  |  |  |  |
| - Fully recovered | 40 (97.6) | 12 (92.3) | 6 (100) | 9 (100) | 13 (100) | NA |
| - Not fully recovered, deteriorated, permanent damage, or death | 0 | 0 | 0 | 0 | 0 |  |
| - Ongoing | 1 (2.4) | 1 (7.7)<sup>†††</sup> | 0 | 0 | 0 |  |
| Action taken by physician, n (%): |  |  |  |  |  |  |
| - None | 6 (14.6) | 1 (7.7) | 0 | 2 (22.2) | 3 (23.1) | NA |
| - Medication given | 35 (85.4) | 12 (92.3) | 6 (100) | 7 (77.8) | 10 (76.9) |  |
| - Study discontinuation | 0 | 0 | 0 | 0 | 0 |  |
| - Hospitalization<sup>‡‡‡</sup> | 0 | 0 | 0 | 0 | 0 |  |

### Preliminary efficacy: microscopy endpoints

In modified ITT analyses, the BV incidence rate by Nugent score during the intervention period was 10.18 per person-year at risk (PY) in the control group, and lower in the metronidazole (1.41/PY; p=0.004), EF+ (3.58/PY; p=0.043), and GynLP groups (5.36/PY; p=0.220) (Table 3). Mean Nugent scores during the intervention period were highest in the control group, lowest in the metronidazole group, and in-between in the two vaginal probiotics groups (Figure 2A). By the end of the intervention period, many women had developed microbiological-BV without symptoms. In line with standard practice, they were not treated, but they were also no longer included in the ‘person-years at risk’ denominator because they had already developed the endpoint. BV incidence rates were therefore much lower (1.26/PY overall) after cessation of the intervention, and similar between groups. The results for BV incidence by modified Amsel criteria were similar to those for Nugent scores (Table 3), and no vulvovaginal candidiasis was diagnosed after randomization.

**Table 3.** Preliminary efficacy – microscopy endpoints (modified ITT analyses)

| BV IR Enr-M2 | Controls |  | Metronidazole |  | EF+ |  | GynLP |  |
| --- | --- | --- | --- | --- | --- | --- | --- | --- |
|  | n/N* | IR (95% CI)<sup>†</sup> | n/N* | IR (95% CI)<sup>†</sup> | n/N* | IR (95% CI)<sup>†</sup> | n/N* | IR (95% CI)<sup>†</sup> |
| Nugent 7-10 | 9/11 | 10.18<br>(5.48 – 18.92) | 2/10 | 1.41<br>(0.35 – 5.62) | 5/12 | 3.58<br>(1.61 – 7.96) | 6/10 | 5.36<br>(2.41 – 11.93) |
| Modified Amsel<sup>‡</sup> | 11/15 | 7.53<br>(4.28 – 13.26) | 3/11 | 2.04<br>(0.66 – 6.31) | 6/13 | 3.36<br>(1.51 – 7.48) | 4/9 | 3.35<br>(1.26 – 8.92) |
| BV IR M2-M6 |  |  |  |  |  |  |  |  |
| Nugent 7-10 | 4/11 | 0.91<br>(0.34 – 2.41) | 5/8 | 1.86<br>(0.78 – 4.48) | 6/9 | 1.58<br>(0.71 – 3.52) | 2/7 | 0.76<br>(0.19 – 3.03) |
| Modified Amsel<sup>‡</sup> | 3/12 | 2.96<br>(0.33 – 3.15) | 5/10 | 2.25<br>(0.94 – 5.40) | 6/8 | 3.84<br>(1.72 – 8.54) | 3/8 | 1.83<br>(0.59 – 5.67) |
| BV IRR Enr-M2 | Controls |  | Metronidazole |  | EF+ |  | GynLP |  |
|  |  |  | IRR (95% CI)<sup>‡</sup> |  | IRR (95% CI)<sup>‡</sup> |  | IRR (95% CI)<sup>‡</sup> |  |
| Nugent 7-10 | NA |  | 0.14 (0.01 – 0.65) |  | 0.35 (0.10 – 1.07) |  | 0.53 (0.16 – 1.60) |  |
| Modified Amsel<sup>‡</sup> | NA |  | 0.27 (0.05 – 1.00) |  | 0.45 (0.14 – 1.28) |  | 0.44 (0.10 – 1.47) |  |
| BV IRR M2-M6 |  |  |  |  |  |  |  |  |
| Nugent 7-10 | NA |  | 2.06 (0.44 – 10.37) |  | 1.74 (0.41 – 8.41) |  | 0.84 (0.08 – 5.85) |  |
| Modified Amsel<sup>‡</sup> | NA |  | 2.22 (0.43 – 14.27) |  | 3.78 (0.81 – 23.35) |  | 1.80 (0.24 – 13.46) |  |

Abbreviations: BV, bacterial vaginosis; CI, confidence interval; EF+, Ecologic Femi+; Enr, enrollment visit; GynLP, Gynophilus LP; IR, incidence rate; IRR, incidence rate ratio; ITT, intent to treat; M2/6, month 2/6 visit; NA, not applicable. There were no cases of vulvovaginal candidiasis during follow-up. Participants who were BV-negative by Amsel but BV-positive by Nugent criteria at the enrollment visit (n=17) were omitted from modified ITT analyses. \*Number of women (n) who developed at least one incident infection during the specified time period as a proportion of the women who had at least one follow-up visit in that time period (N). <sup>†</sup>Incident infections divided by person-years at risk. Three participants in the Enr-M2 BV by modified Amsel model, seven in the Enr – M2 BV by Nugent model, 10 in the M2-M6 BV by modified Amsel model, and 11 in the M2-M6 BV by Nugent model were omitted due to having been at risk for less than 10 person-days. <sup>‡</sup>The modified Amsel criteria are positive when at least two of the three laboratory criteria are positive: vaginal pH>4.5, whiff test positive, and/or presence of >20% clue cells on wet mount. <sup>‡</sup>Compared to the control group.

**Figure 2.**
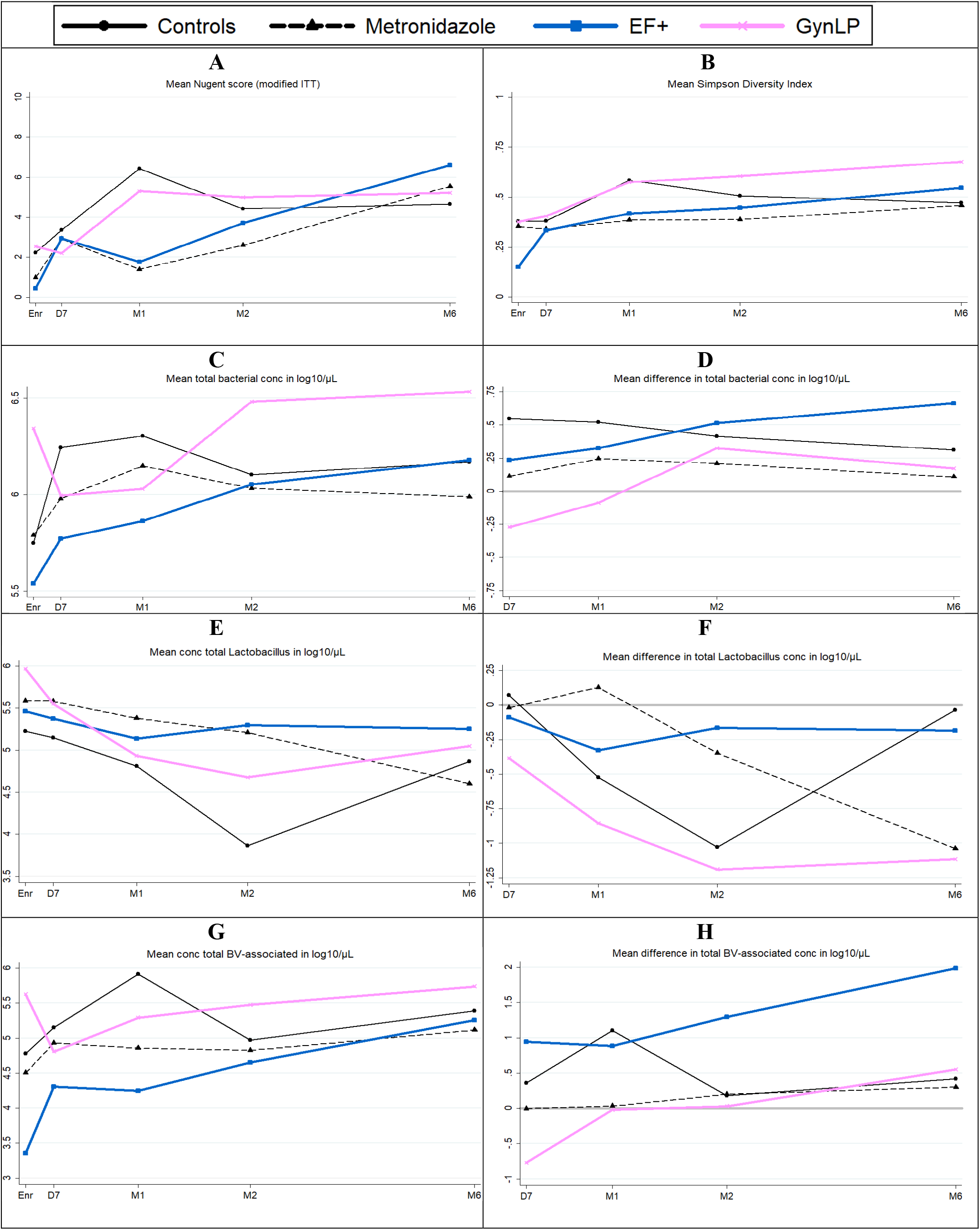
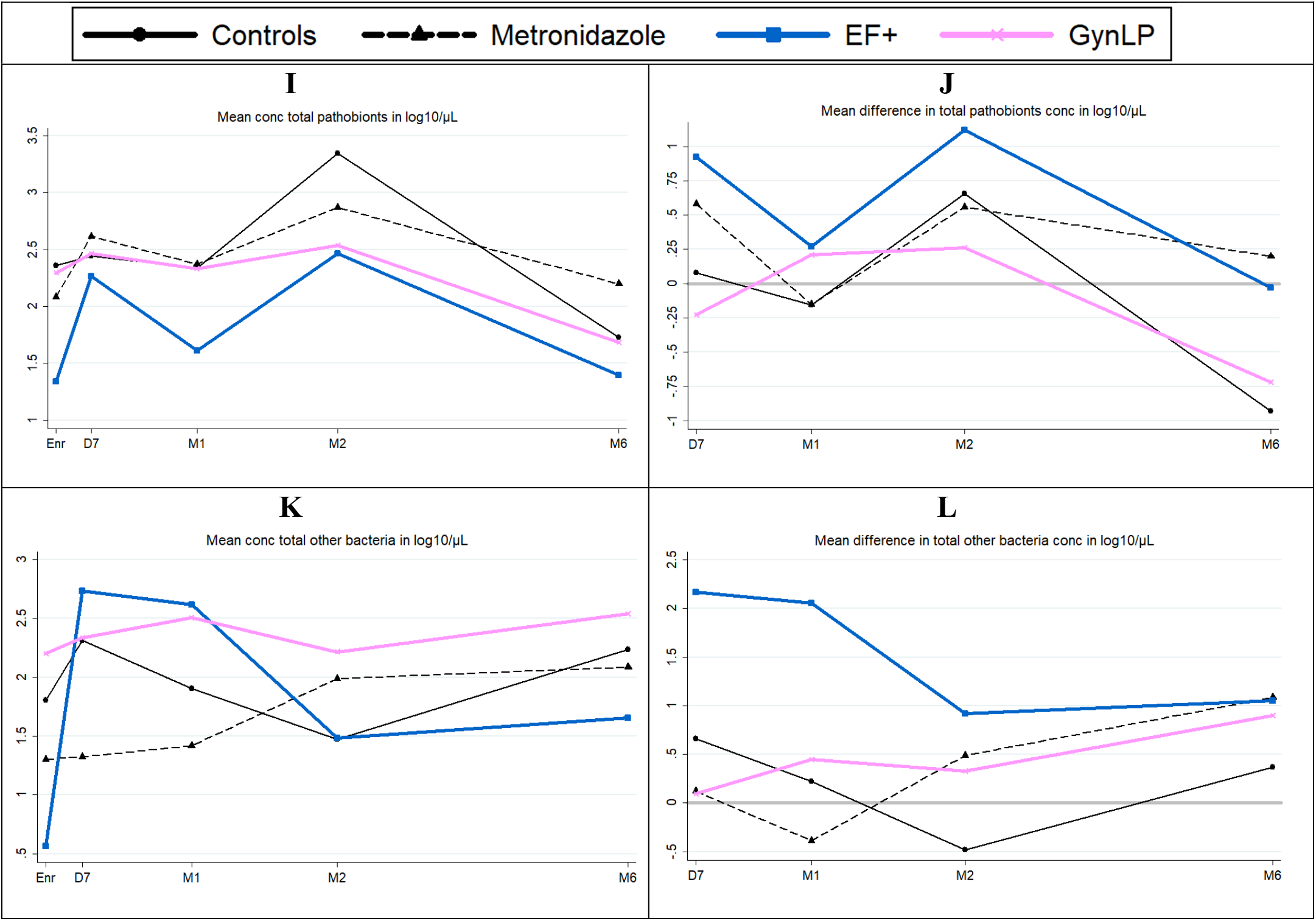
Preliminary efficacy by Nugent score, alpha diversity, and bacterial group concentration. Abbreviations: Conc, concentration; D7, Day 7 visit; EF+, Ecologic Femi+; Enr, enrollment visit; GynLP, Gynophilus LP; ITT, intention to treat; M1/2/6, month 1/2/6 visit; Scr, screening visit; VMB, vaginal microbiota. Changes in VMB outcomes over time per randomization group. See Supplement Table S4 for 95% confidence intervals. Mean Nugent scores over time, only including women (n=51) who were BV-negative by Amsel and Nugent criteria (modified ITT analysis). (**B**) Mean alpha diversity over time. (**C**) Mean bacterial cell concentration over time. (**D**) Difference in mean bacterial cell concentration with enrollment, over time. (**E**) Mean lactobacilli concentration over time. (**F**) Difference in mean lactobacilli concentration with enrollment, over time. (**G**) Mean BV-associated anaerobes concentration over time. (**H**) Difference in mean BV-associated anaerobes concentration with enrollment, over time. (**I**) Mean pathobionts concentration over time. (**J**) Difference in mean pathobionts concentration with enrollment, over time. (**K**) Mean other bacteria concentration over time. (**L**) Difference in mean other bacteria concentration with enrollment, over time.

### Preliminary efficacy: sequencing endpoints

We took the baseline VMB composition imbalances into account by not only showing bacterial group means at each visit but also mean differences with enrollment (Figure 2 and Table S4 for concentrations, and Figure S3 and Table S5 for relative abundances), and by using mixed effects models including participant identification number as the random effect to determine if any changes were statistically significant (Table 4). Immediately after BV treatment completion, the VMBs of most women gradually worsened (lactobacilli declined and BV-anaerobes expanded) due to the high-risk nature of the cohort. The mean lactobacilli concentration declined to a low of 3.86 log_10_ cells/μl at M2 in the control group, but less so in the intervention groups (ranging from 4.60-5.58 log_10_ cells/μl at follow-up visits). In unadjusted mixed effects models using data from the intervention period only, metronidazole users had a higher lactobacilli concentration (p=0.043) and relative abundance (p=0.006) than controls, EF+ users had a higher relative abundance (p=0.014) but not concentration than controls, and GynLP users had trends in the same directions that were not statistically significant. The expansion of BV-anaerobes was significantly lower in oral metronidazole users (relative abundance; p=0.023), and in EF+ users (concentration; p=0.041), compared to controls. Mean pathobionts concentrations were low in all groups throughout, ranging from 1.61-3.35 log_10_ cells/μl at follow-up visits. Mixed effects models did not identify any significant associations between randomization groups and pathobionts concentrations or relative abundances, but showed trends (0.05<p<0.1) towards lower pathobionts relative abundances in the two vaginal probiotics groups compared to controls (Table 4). Mean concentrations of ‘other bacteria’ were also low throughout, but highest in the EF+ group at the D7 and M1 visits (EF+ contains a *Bifidobacterium* strain). This was significant in unadjusted mixed effects models for relative abundances (p=0.023) but not concentrations.

**Table 4.** Preliminary efficacy – mixed effects models.

| Unadjusted mixed effects models | Metro |  | EF+ |  | GynLP |  |
| --- | --- | --- | --- | --- | --- | --- |
|  | OR (95% CI)* | p* | OR (95% CI)* | p* | OR (95% CI)* | p* |
| Nugent score<sup>†</sup> | 0.16 (0.02 – 1.09) | 0.062 | 0.25 (0.04 – 1.66) | 0.151 | 1.68 (0.24 – 11.56) | 0.599 |
| Nugent score categories<sup>†</sup> |  |  |  |  |  |  |
| - 4-6 vs 0-3 | 1.04 (0.20 – 5.50) | 0.960 | 0.36 (0.05 – 2.43) | 0.293 | 2.16 (0.38 – 12.31) | 0.387 |
| - 7-10 vs 0-3 | 0.08 (0.01 – 0.80) | 0.032 | 2.16 (0.38 – 12.31) | 0.148 | 1.12 (0.15 – 8.28) | 0.913 |
| Total bacterial conc<sup>†</sup> | 0.85 (0.53 – 1.36) | 0.497 | 0.72 (0.45 – 1.14) | 0.160 | 0.96 (0.60 – 1.55) | 0.877 |
| Total *Lactobacillus* conc<sup>†</sup> | 2.14 (1.02 – 4.49) | 0.043 | 1.86 (0.90 – 3.86) | 0.095 | 1.43 (0.67 – 3.04) | 0.352 |
| Total BV-associated conc<sup>†</sup> | 0.58 (0.23 – 1.49) | 0.260 | 0.38 (0.15 – 0.96) | 0.041 | 0.89 (0.34 – 2.31) | 0.812 |
| Total pathobionts conc<sup>†</sup> | 0.91 (0.30 – 2.75) | 0.865 | 0.57 (0.19 – 1.71) | 0.318 | 0.79 (0.25 – 2.43) | 0.676 |
| Total other bacteria conc<sup>†</sup> | 0.72 (0.27 – 1.90) | 0.507 | 1.42 (0.55 – 3.70) | 0.471 | 1.60 (0.60 – 4.29) | 0.349 |
| Total *Lactobacillus* RA<sup>‡</sup> | 1.36 (1.09 – 1.70) | 0.006 | 1.32 (1.06 – 1.64) | 0.014 | 1.10 (0.88 – 1.38) | 0.408 |
| Total BV-associated RA<sup>‡</sup> | 0.79 (0.65 – 0.97) | 0.023 | 0.83 (0.68 – 1.01) | 0.067 | 1.00 (0.81 – 1.22) | 0.973 |
| Total pathobionts RA<sup>‡</sup> | 0.93 (0.84 – 1.03) | 0.159 | 0.92 (0.83 – 1.01) | 0.093 | 0.92 (0.83 – 1.02) | 0.100 |
| Total other bacteria RA<sup>‡</sup> | 1.00 (1.00 – 1.01) | 0.739 | 1.01 (1.00 – 1.01) | 0.023 | 1.00 (1.00 – 1.01) | 0.671 |
| Pooled VMB type<sup>‡</sup> |  |  |  |  |  |  |
| - LA vs LD | 1.17 (0.24 – 5.78) | 0.849 | 1.20 (0.24 – 5.92) | 0.823 | 1.72 (0.33 – 9.04) | 0.520 |
| - BV vs LD | 0.02 (0.00 – 0.43) | 0.012 | 0.04 (0.00 – 0.73) | 0.029 | 0.39 (0.03 – 5.50) | 0.487 |
| - PB vs LD | 0.06 (0.00 – 1.54) | 0.090 | 0.08 (0.00 – 1.77) | 0.109 | 0.14 (0.01 – 3.34) | 0.225 |
| Simpson diversity<sup>‡</sup> | 0.90 (0.77 – 1.06) | 0.200 | 0.94 (0.80 – 1.10) | 0.454 | 1.07 (0.91 – 1.26) | 0.429 |
| **Adjusted mixed effects models<sup>§</sup>** |  |  |  |  |  |  |
| Nugent score<sup>†</sup> | 0.19 (0.03 – 1.31) | 0.092 | 0.26 (0.04 – 1.85) | 0.178 | 2.05 (0.30 – 13.92) | 0.464 |
| Nugent score categories<sup>†</sup> |  |  |  |  |  |  |
| - 4-6 vs 0-3 | 1.24 (0.20 – 7.82) | 0.821 | 0.36 (0.04 – 3.13) | 0.353 | 2.42 (0.36 – 16.26) | 0.362 |
| - 7-10 vs 0-3 | 0.06 (0.00 – 0.77) | 0.031 | 0.19 (0.02 – 1.90) | 0.156 | 1.42 (0.17 – 11.81) | 0.747 |
| Total bacterial conc<sup>†</sup> | 0.75 (0.47 – 1.18) | 0.208 | 0.64 (0.40 – 1.02) | 0.061 | 0.90 (0.57 – 1.43) | 0.666 |
| Total *Lactobacillus* conc<sup>†</sup> | 1.74 (0.83 – 3.67) | 0.142 | 1.47 (0.69 – 3.16) | 0.319 | 1.26 (0.60 – 2.68) | 0.541 |
| Total BV-associated conc<sup>†</sup> | 0.57 (0.22 – 1.47) | 0.243 | 0.37 (0.14 – 0.98) | 0.046 | 0.91 (0.35 – 2.36) | 0.848 |
| Total pathobionts conc<sup>†</sup> | 0.99 (0.35 – 2.77) | 0.980 | 0.66 (0.23 – 1.81) | 0.445 | 0.98 (0.36 – 2.90) | 0.965 |
| Total other bacteria conc<sup>†</sup> | 0.65 (0.25 – 1.69) | 0.374 | 1.31 (0.49 – 3.50) | 0.591 | 1.79 (0.68 – 4.71) | 0.239 |
| Total *Lactobacillus* RA<sup>‡</sup> | 1.32 (1.06 – 1.65) | 0.014 | 1.30 (1.03 – 1.64) | 0.025 | 1.06 (0.85 – 1.33) | 0.601 |
| Total BV-associated RA<sup>‡</sup> | 0.81 (0.66 – 1.00) | 0.049 | 0.83 (0.67 – 1.03) | 0.098 | 1.02 (0.83 – 1.26) | 0.855 |
| Total pathobionts RA<sup>‡</sup> | 0.93 (0.84 – 1.03) | 0.180 | 0.92 (0.83 – 1.02) | 0.120 | 0.93 (0.84 – 1.03) | 0.147 |
| Total other bacteria RA<sup>‡</sup> | 1.00 (1.00 – 1.01) | 0.436 | 1.01 (1.00 – 1.01) | 0.009 | 1.00 (1.00 – 1.01) | 0.439 |
| Pooled VMB type<sup>‡</sup> |  |  |  |  |  |  |
| - LA vs LD | 1.46 (0.26 – 8.11) | 0.665 | 1.59 (0.27 – 9.55) | 0.610 | 2.11 (0.37 – 12.15) | 0.401 |
| - BV vs LD | 0.02 (0.00 – 0.48) | 0.017 | 0.02 (0.00 – 0.64) | 0.027 | 0.44 (0.02 – 7.84) | 0.575 |
| - PB vs LD | 0.08 (0.00 – 1.49) | 0.090 | 0.11 (0.01 – 2.02) | 0.136 | 0.21 (0.01 – 3.71) | 0.284 |
| Simpson diversity<sup>‡</sup> | 0.93 (0.78 – 1.09) | 0.359 | 0.96 (0.81 – 1.13) | 0.597 | 1.10 (0.93 – 1.30) | 0.272 |

Abbreviations: BV, bacterial vaginosis; CI, confidence interval; conc, concentration; EF+, Ecologic Femi+; Enr, enrollment visit; GynLP, Gynophilus LP; LA, lactobacilli and anaerobes VMB type; LD, *Lactobacillus*-dominated VMB type; Metro, metronidazole group; OR, odds ratio; PB, pathobionts VMB type; VMB, vaginal microbiota. \*Compared to the control group. †Including all valid samples during product use (D7, M1, and M2 visits). Self-sampled samples were also taken during product use, but were not Gram stained nor tested by 16S rRNA gene qPCR (see Methods). ‡Including all valid samples during product use (D7, M1, and M2 visits, and self-sampled samples). §Model adjusted for hormonal/pregnancy status, sexual risk taking, and age (see Methods).

The proportions of women in each group at each visit having a particular VMB type corresponded with the concentration and relative abundance data, and additionally showed that – in lactobacilli-dominated women – the Li VMB type was far more common than the Lcr and Lo VMB types throughout (Figure S4). Among women with dysbiosis, the LA and BV_GV VMB types continued to be the most common dysbiosis types during follow-up. In unadjusted mixed effects models using intervention period data only, metronidazole users and EF+ users, each compared to controls, were significantly less likely to have dysbiotic VMB types (BV_GV, BV_noGV, and GV combined; p=0.012 and p=0.029, respectively) (Table 4).

The associations in unadjusted mixed effects models persisted after adjustment for hormonal contraception use/pregnancy, sexual risk taking, and age, except for the association with *Lactobacillus* concentration among metronidazole users. Metronidazole users compared to controls had a significantly higher *Lactobacillus* relative abundance (p=0.014), a significantly lower BV-associated bacteria relative abundance (p=0.049), and were significantly less likely to have BV by Nugent scoring (p=0.031) or by VMB types (BV_GV, BV_noGV, and GV combined; p=0.017) (Table 4). EF+ users compared to controls had a significantly higher *Lactobacillus* relative abundance (p=0.025), a significantly lower BV-associated bacteria concentration (p=0.046), a significantly higher relative abundance of ‘other bacteria’ (p=0.009), and were significantly less likely to have BV-like VMB types (p=0.027).

### Detection of probiotic strains

During the intervention period, relevant probiotic strains were detected in 39% of samples from EF+ users and 20% of samples from GynLP users (all swabs combined, including self-sampled swabs). The detection percentages were 58% and 31%, respectively, in sensitivity analyses using non-rarefied data. Some of the EF+ strains cannot be differentiated from naturally occurring strains, and EF+-like strains were therefore detected (at low levels) in all groups at most time points (Figure 3, Table S4). However, the mean concentrations were highest in the EF+ group during the intervention period (mean concentrations 0.48-1.92 log_10_ cells/μl per visit for all women combined, and 3.62-4.28 log_10_ cells/μl per visit for women who did have EF+ strains detected using rarefied data). The GynLP strain was only detected in the GynLP group during the intervention period (mean concentrations 0.25-1.05 log_10_ cells/μl per visit for all women combined, and 3.72-4.55 log_10_ cells/μl per visit for women who did have GynLP detected using rarefied data). Inter- and intra-individual differences between participants were high: the highest vaginal probiotic concentration detected in an individual EF+ user was 5.51 log_10_ cells/μl, and in an individual GynLP user was 6.17 log_10_ cells/μl. During the intervention period, the mean relative abundances of the probiotic strains were 0.03 in both EF+ and GynLP users, and 0.08 and 0.15, respectively, if only samples in which any strains were detected were included.

**Figure 3.**
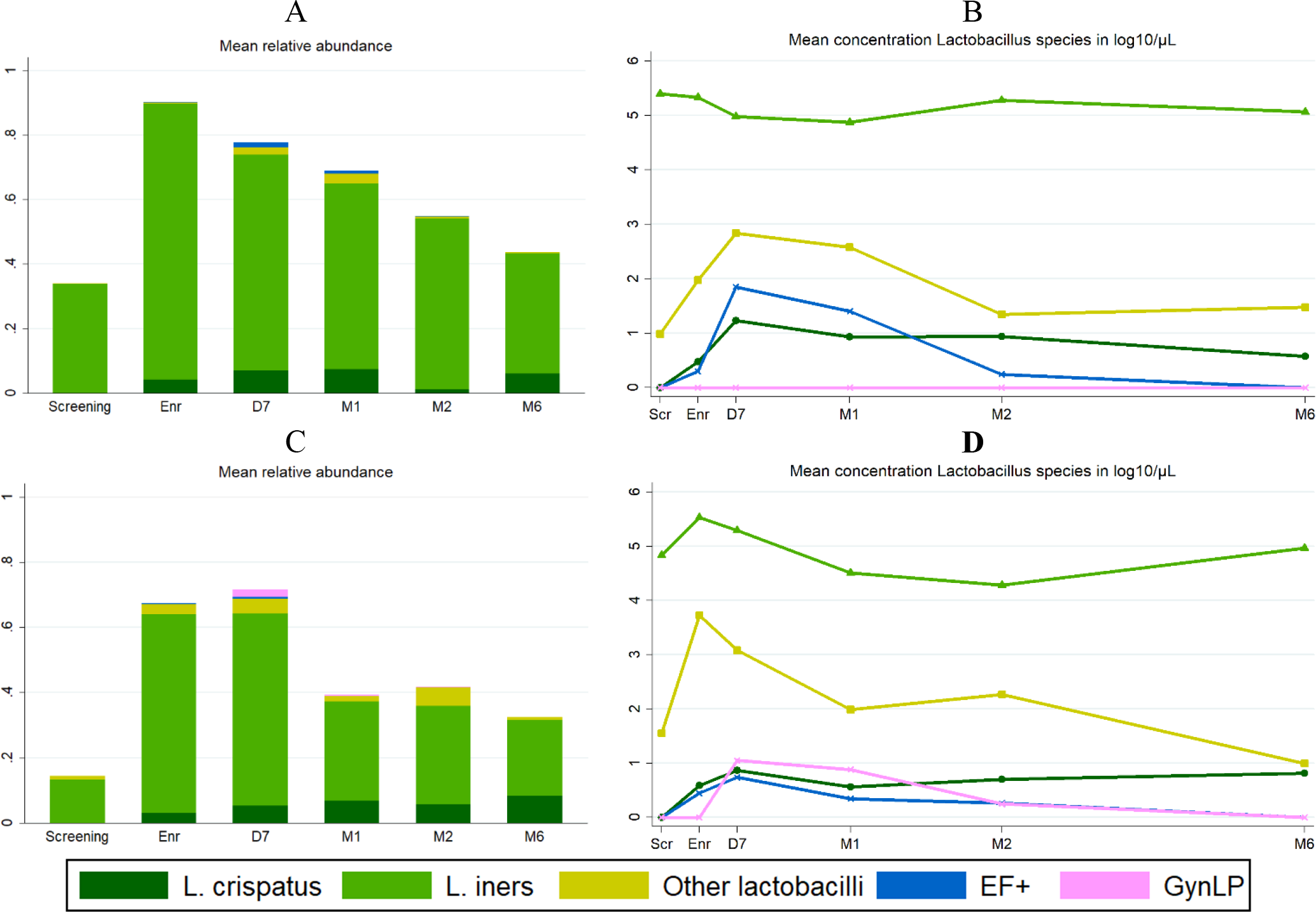
Detection of probiotic strains during the trial. Abbreviations: D7, Day 7 visit; EF+, Ecologic Femi+; Enr, enrollment visit; GynLP, Gynophilus LP; M1/2/6, month 1/2/6 visit; Scr, screening visit. Mean relative abundance **(A)** and mean concentration **(B)** of *Lactobacillus* species over time in the EF+ group; mean relative abundance **(C)** and mean concentration **(D)** of *Lactobacillus* species over time in the GynLP group. The length of bars in (A) and (C) depicts total relative abundance of all *Lactobacillus* species combined.

### VMB transitions

The stacked graph and alluvial diagrams (Figure S4) show that transitions from one VMB type to another were common. As expected, most transitions during the intervention period were from *Lactobacillus*-dominated states to dysbiotic states, but the reverse also occurred. Transitions between VMB types were common in all randomization groups. The percentage of actual transitions divided by potential transitions between D7 and M6 were 29/66 (43.9%) in the control group, 32/64 (50.0%) in the metronidazole group, 28/67 (41.8%) in the EF+ group and 24/56 (42.9%) in the GynLP group (Fisher’s exact p=0.792).

### Incidence of sexually transmitted and urinary tract infections

As expected, the incidences of HIV (n=2), herpes simplex type 2 (n=1), syphilis (n=1), gonorrhea (n=5), chlamydia (n=6), TV (n=5) and urinary tract infection (n=7) were too low to determine differences between randomization groups (Table S7).

## DISCUSSION

This trial showed that all three interventions were safe. Our preliminary efficacy results confirm that intermittent use of metronidazole reduces BV recurrence^10–12^, and suggest that intermittent use of lactobacilli-containing vaginal probiotics may also reduce BV recurrence. We also found that vaginal probiotic use is acceptable and feasible in African settings (to be reported elsewhere). These findings are important because many women and clinicians would prefer safe and efficacious vaginal probiotics to metronidazole as they are not expected to negatively affect other body niche microbiotas or cause antimicrobial resistance, and may have fewer side effects.

Our trial was funded as a pilot study and therefore had a modest sample size. Despite this, many of the preliminary efficacy associations for intermittent metronidazole and EF+ use reached statistical significance. While this was not the case for intermittent GynLP use, all of the trends in this group were in the same directions. We believe that the GynLP group suffered a few disadvantages compared to the other randomization groups, which may explain the lack of statistical significance. Randomization imbalances commonly occur in small trials^36^, and in our trial, this led to women in the GynLP group being more dysbiotic at baseline than controls. We ameliorated this disadvantage in our analysis strategy, but we may not have been able to eradicate it. In addition, GynLP users were less adherent on average than metronidazole and EF+ users. Differences in adherence using triangulated data did not reach statistical significance, but probiotic strain detection rates (58% for EF+ samples and 31% for GynLP samples using non-rarefied data) support this claim. Biose has since simplified the dosing regimen of next generation Gynophilus products to two fixed days a week to boost adherence. Finally, GynLP dosing (once every four days) was similar to metronidazole dosing (twice weekly) but less frequent than EF+ dosing (once per day for the first five days followed by thrice weekly for the remainder of the intervention period).

Probiotic detection rates were 58% for EF+ samples and 31% for GynLP samples, and inter- and intra-individual variabilities were high (with probiotic concentrations ranging from zero to 6.17 log_10_ cells/μl). This detection variability is consistent with most other vaginal probiotic studies that used sampling at non-daily intervals and molecular assessment methods^21,37–39^. A major drawback of all of those studies, including ours, is that product use was not directly observed but self-reported, and precise information about the time period between last product insertion and sample collection was lacking. The average total bacterial load of a healthy vagina is currently not known^40^. The average vaginal surface area was estimated to be 87.5 cm^41^. One Dacron swab head in this study absorbed about 10^6^/μL bacteria. If we assume that one swab head absorbs on average 200 μL^42^, and that this covers about one cm^2^ of a total of about 100 cm^2^ vaginal surface, the total bacterial load in the vagina would be in the order of 2×10^10^ bacteria. The vaginal probiotic strain(s) in our trial were both applied at about 1.5×10^9^ CFU per dose, which would be about 7.5% of the total vaginal load after application if all probiotic bacteria were to remain in the vagina. We detected mean relative abundances of 7.7% for EF+ strains and 15.1% for GynLP when only samples with any relevant probiotic strains detected during product use were included (thereby eliminating any potential non-adherence). A recently published study of Gynophilus Slow Release tablet (which is almost identical to GynLP) in which women self-sampled every day showed that mean vaginal concentrations of Lcr35 by qPCR were between 10^4^ and 10^6^ CFU/μl in women who used the tablet once every four or five days^43^. Using our estimated concentration data, we detected a similar concentration range in samples with any GynLP detected. This consistency is reassuring, but the question remains whether probiotic concentrations of this order of magnitude optimally prevent BV recurrence in the long-term. Furthermore, all studies referenced in this paragraph, including ours, have shown that probiotic strains do not persist in the vagina after dosing has ceased. The second question then is whether the colonization capacity of probiotic bacteria should be improved. Our data suggest that probiotic lactobacilli may boost ‘natural’ lactobacilli indirectly, which may be sufficient. Indirect effects may include increased localized lactic acid production, modulation of cervicovaginal mucosal immune responses, and/or inhibition of biofilm formation, by probiotic bacteria^3,44,45^.

Additional limitations of our study include the high urogenital infection risk of this cohort (which makes prevention more challenging), and our inability to fully control for potential confounders. However, the mixed effects models were controlled for some of the best known VMB determinants (hormonal contraception, pregnancy, sexual risk taking, and age)^1,46–48^. We were not able to exclude women with gonorrhea and/or chlamydia infection at the time of randomization due to the slow laboratory turn-around time, but the VMB compositions of women with and without infection were similar, and we therefore think that this did not negatively affect our results.

With the development of better genomic and culturing methods, we are now on the cusp of a new era in vaginal probiotic research. Past vaginal probiotic studies have shown mixed results^13–24^, but almost all of these studies used imprecise VMB assessments based on clinical symptoms and microscopy. The addition of sequencing methods showed that many more women than previously thought are not lactobacilli-dominated after standard antibiotic BV treatment, that host responses to antibiotic and probiotic treatment are highly variable, and that it is possible to differentiate between probiotic strains and ‘natural’ lactobacilli. Furthermore, others have shown that quantifying relative abundance data in the same manner as we have done in this study correlates well with species-specific quantitative PCRs of non-minority species^33,34^. This then allows for microbiota data reduction into quantitative variables that can be analyzed in mixed effects models that adjust for repeated measures and confounding. We recommend that future trials incorporate these or other rigorous methods, optimize dosing and timing of product insertion versus sample collection, and enroll women with various urogenital risk profiles. Ideally, these trials would also evaluate the effects of interventions on vaginal biofilm formation, and – eventually – the impact on pregnancy complications, HIV epidemics, and other adverse outcomes.

## Data Availability

All data will be made available via the University of Liverpool Data Research Catalogue (https://datacat.liverpool.ac.uk/) with publication of the manuscript in a peer-reviewed journal. We prefer to do this after peer-review because the peer-review may result in additional analyses. The most essential data are included in the manuscript and in Supplements 1 and 2.

## Acknowledgments

We thank the study participants, the Rinda Ubuzima team, the National Reference Laboratory in Kigali, the Centre for Genomic Research and research support staff at the University of Liverpool, Christina Gill, Robert Meester, Mike Humphreys, Jessica Younes, Vicky Jespers, Tania Crucitti, and Anna Maria Geretti. This work was funded by the DFID/MRC/Wellcome Trust Joint Global Health Trials Scheme as a Development Project (grant reference MR/M017443/1; grant title: “Preparing for a clinical trial of interventions to maintain normal vaginal microbiota for preventing adverse reproductive health outcomes in Africa”) and the University of Liverpool (Technology Directorate Voucher). Vaginal probiotics for use in the trial were donated free of charge by Winclove Probiotics (Amsterdam, The Netherlands) and Biose (formerly Probionov, Aurillac, France). The findings and conclusions in this paper are those of the authors and do not necessarily represent the views of the authors’ institutions or companies, or the funders.

## Authors’ contributions

JvdW obtained the research funding and wrote the study protocol and data collection documents. JvdW, AN, EL, and JR were members of the Trial Steering Committee. SA, LM, VM, MU, and JvdW collected the primary data. LM and VM performed the diagnostic testing. MV, CB, AD, and JR conducted or oversaw the 16S rRNA gene sequencing and qPCRs. JvdW and MV developed the analytical approach and performed the statistical analyses. JvdW and MV wrote the manuscript. All authors commented on and approved the final manuscript. JvdW had full access to the data and had final responsibility for the decision to submit for publication.

